# Casirivimab and Imdevimab Treatment in Seropositive, Hospitalized COVID-19 Patients With Non-neutralizing or Borderline Neutralizing Antibodies

**DOI:** 10.1101/2022.06.14.22276389

**Authors:** Andrea T. Hooper, Selin Somersan-Karakaya, Shane E. McCarthy, Eleftherios Mylonakis, Shazia Ali, Jingning Mei, Rafia Bhore, Adnan Mahmood, Gregory P. Geba, Paula Dakin, David M. Weinreich, George D. Yancopoulos, Gary A. Herman, Jennifer D. Hamilton, the COVID-19 Phase 2/3 Hospitalized Trial Team

## Abstract

We conducted a post-hoc analysis in seropositive patients who were negative or borderline for functional neutralizing antibodies (nAbs) against SARS-CoV-2 at baseline from a phase 1/2/3 trial of casirivimab and imdevimab (CAS+IMD) treatment in hospitalized COVID-19 patients on low-flow or no supplemental oxygen prior to the emergence of Omicron-lineage variants. Patients were randomized to a single dose of 2.4 g CAS+IMD, 8.0 g CAS+IMD, or placebo. Patients seropositive for anti-SARS-CoV-2 antibodies at baseline were analyzed by their baseline nAb status. At baseline, 20.6% (178/864) of seropositive patients were negative/borderline for nAbs. CAS+IMD reduced viral load in patients who were negative/borderline for nAbs versus placebo, but not in patients who were positive for nAbs. We observed a trend in reduction of the proportion of patients who died or required mechanical ventilation (MV), as well as in all-cause mortality, by day 29 with CAS+IMD versus placebo in patients who were negative/borderline for nAbs. In those who were negative/borderline for nAbs, the proportions who died/needed MV from days 1–29 were 19.1% and 10.9%, and the proportions of patients who died from days 1–29 were 16.2% and 9.1%, in the placebo and CAS+IMD combined dose groups, respectively. No measurable harm or benefit in death/MV or all-cause mortality was observed in patients who were positive for nAbs. In hospitalized COVID-19 patients on low-flow or no supplemental oxygen, CAS+IMD reduced viral load, the risk of death or MV, and all-cause mortality in seropositive patients who were negative/borderline for nAbs.

**Importance:** The clinical benefit of CAS+IMD in hospitalized seronegative patients with COVID-19 has previously been demonstrated, although these studies observed no clinical benefit in seropositive patients. As the prevalence of SARS-CoV-2 seropositive individuals rises due to both vaccination and previous infection, it is important to understand whether there is a subset of hospitalized patients with COVID-19, who have antibodies against SARS-CoV-2, who could benefit from anti-SARS-CoV-2 monoclonal antibody treatment. This post-hoc analysis demonstrates that there is a subset of hospitalized, seropositive patients with inadequate SARS-CoV-2 nAbs (ie, those who were negative or borderline for nAbs) who may still benefit from CAS+IMD treatment if infected with a susceptible variant. Therefore, utilizing seronegativity status alone to guide treatment decisions for patients with COVID-19 may fail to identify seropositive patients who could benefit from anti-SARS-CoV-2 monoclonal antibody therapies which retain activity against circulating strains, depending on how effectively their endogenous antibodies neutralize SARS-CoV-2.

## INTRODUCTION

Severe acute respiratory syndrome coronavirus 2 (SARS-CoV-2) continues to evolve, and data have shown that complete coronavirus disease 2019 (COVID-19) vaccination with a booster is protective against symptomatic disease and severe COVID-19 (1–3). As the prevalence of SARS-CoV-2 seropositive individuals rises due to both vaccination and previous infection (4), it is important to understand whether there is a subset of hospitalized patients with COVID-19, who have antibodies against SARS-CoV-2, who could benefit from anti-SARS-CoV-2 monoclonal antibody treatment.

Monoclonal antibody therapeutics benefit patients across the spectrum of COVID-19 disease severity (5), as well as individuals who can neither receive nor respond to vaccines (6, 7). Casirivimab and imdevimab (CAS+IMD) is a combination of two neutralizing monoclonal antibodies that bind non-overlapping epitopes of the SARS-CoV-2 spike protein receptor-binding domain (8). Prior to the emergence of the Omicron variant, CAS+IMD was shown to be effective in the treatment of outpatients with COVID-19 (9) – reducing viral load, decreasing risk of hospitalization or death, and decreasing symptom duration – as well as demonstrating efficacy in the prevention of COVID-19 (10).

The efficacy and safety of CAS+IMD in hospitalized patients with COVID-19 was demonstrated in an open-label platform trial in the UK (RECOVERY) (11) as well as a phase 1/2/3, double-blinded, placebo-controlled trial in patients on low-flow or no supplemental oxygen (Study 2066) (12). In the RECOVERY study, CAS+IMD reduced mortality in hospitalized patients with COVID-19, but this benefit was observed only in patients who were seronegative (ie, had no measurable antibody immunity to SARS-CoV-2) at baseline (11). Consistent with these data, clinical benefit, including improvements in death or need for mechanical ventilation, all-cause mortality, and discharge from hospital, were observed in the overall population in Study 2066, driven by the benefit in seronegative patients and with no benefit or harm observed in seropositive patients (12).

In the current landscape, where the majority of the population is vaccinated against COVID-19 and/or has a history of SARS-CoV-2 infection, the estimated anti-SARS-CoV-2 seropositivity rate in US is 94.7% (13). Thus, it is important to better understand the potential role of CAS+IMD treatment in seropositive patients. Although seropositive patients have detectable anti-SARS-CoV-2 antibodies, it was hypothesized that the neutralizing function of these antibodies in a subset of hospitalized patients may be impaired. We questioned whether treatment with CAS+IMD may also provide clinical benefit in certain subsets of seropositive, hospitalized patients with COVID-19. To further investigate this question, we conducted a post-hoc analysis of seropositive patients from Study 2066 who were negative or borderline at baseline for neutralizing antibodies against SARS-CoV-2 as compared with those patients with measurable baseline neutralizing activity. While CAS+IMD has markedly diminished neutralization against the Omicron variant (14), and is not currently authorized in any geographic regions where infection is likely to have been caused by a non-susceptible SARS-CoV-2 variant (15), Study 2066 was conducted prior to the emergence of Omicron and subsequent variants and therefore allowed us to evaluate the benefit of CAS+IMD in a subset of hospitalized patients with susceptible strains of SARS-CoV-2.

## RESULTS

### Patient characteristics by neutralizing antibody status

As of April 9, 2021, prior to the emergence of Delta or Omicron-lineage variants, 2053 patients from phases 1/2/3 on low-flow or no supplemental oxygen were randomized into the study, of whom 2007 were treated and included in the full analysis set (FAS; **Fig. S1**). Of those, 1759 tested positive for SARS-CoV-2 by central lab evaluation, constituting the modified full analysis set (mFAS). Of these patients, a total of 864 (49.1%) were seropositive at baseline (seropositive mFAS), the primary population for the presented analysis: 560 patients in the CAS+IMD combined dose group and 304 patients in the placebo group. Anti-SARS-CoV-2 seropositivity was defined using a composite of three individual assays, as detailed in methods section.

Although antibodies against SARS-CoV-2 are detected in seropositive individuals in this study, the neutralizing function of those antibodies is not characterized by the three serology assays. Therefore, neutralizing antibody status was determined using a high-throughput clinical test that measures the capacity of patient serum samples to neutralize recombinant vesicular stomatitis virus (VSV) encoding the SARS-CoV-2 spike glycoprotein (16). Using this assay, seropositive patients were characterized as positive, negative, or borderline for SARS-CoV-2 functional neutralizing antibodies. At baseline, 20.6% (178/864) of seropositive patients (seropositive mFAS) were negative or borderline for neutralizing antibodies: 110 patients in the CAS+IMD combined dose group and 68 patients in the placebo group. Further serological characterization of seropositive patients by neutralizing antibody status is presented in **Table S1** and **Table S2**.

Baseline demographics and characteristics differed slightly among those who were negative or borderline for neutralizing antibodies as compared with those who were positive for neutralizing antibodies. Patients who were negative or borderline for neutralizing antibodies were older (64.5 versus 60.0 years), had higher baseline viral loads (median values of 6.5 versus 5.2 log_10_ copies/mL), included fewer Hispanic or Latino patients (24.2% versus 39.8%), and had a greater proportion of immunocompromised patients (24.7% versus 12.9%) compared with patients who were positive for neutralizing antibodies, respectively (**Table 1**). Furthermore, seropositive patients with lower neutralizing titers at baseline exhibited higher baseline viral loads (**Fig. 1**).

**FIG 1.**
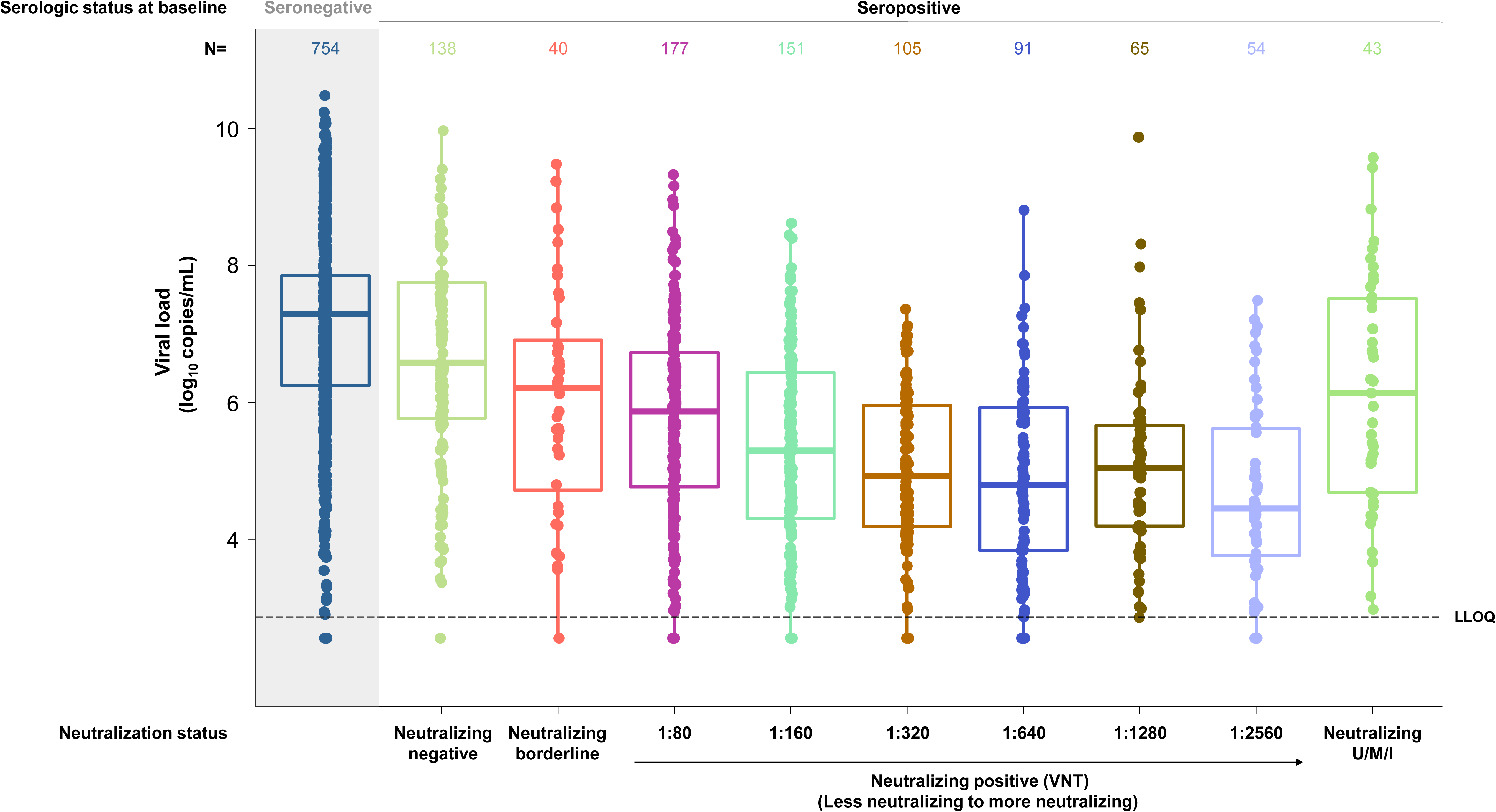
Viral load by baseline neutralizing antibody titer. Serostatus was determined using composite serostatus based on EUROIMMUN anti-spike [S1] IgA and IgG assays and the Abbott anti-nucleocapsid IgG assay. Seropositive is defined as positive in any test; seronegative is defined as negative in all available tests. Neutralizing titer was determined by IMMUNO-COV neutralization assay in seropositive patients only; the seronegative group was not tested in the neutralizing assay. Dots represent individual patient data; boxes represent median and interquartile range. The LLOQ was 2.85 log_10_ copies/mL. mFAS is presented. Ig, immunoglobulin; LLOQ, lower limit of quantification; mFAS, modified full analysis set; VNT, viral neutralizing titer; U/M/I, unknown/missing/indeterminant.

**TABLE 1.** Demographics and baseline characteristics in seropositive patients by baseline neutralizing antibody status^a^

|  | Placebo | CAS+IMD<br>2.4 g IV | CAS+IMD<br>8.0 g IV | CAS+IMD<br>combined doses | Total |
| --- | --- | --- | --- | --- | --- |
| **Baseline neutralizing antibody status: Negative or**<br>**borderline** | ***n* = 68** | ***n* = 57** | ***n* = 53** | ***n* = 110** | ***n* = 178** |
| Age, yr |  |  |  |  |  |
| Median (range) | 66.0 (24:100) | 61.0 (35:94) | 63.0 (37:95) | 62.5 (35:95) | 64.5 (24:100) |
| ≥65, *n* (%) | 40 (58.8) | 26 (45.6) | 23 (43.4) | 49 (44.5) | 89 (50.0) |
| Male sex, *n* (%) | 38 (55.9) | 31 (54.4) | 28 (52.8) | 59 (53.6) | 97 (54.5) |
| Race, *n* (%) |  |  |  |  |  |
| White | 50 (73.5) | 35 (61.4) | 43 (81.1) | 78 (70.9) | 128 (71.9) |
| Black or African American | 8 (11.8) | 8 (14.0) | 4 (7.5) | 12 (10.9) | 20 (11.2) |
| Asian | 2 (2.9) | 3 (5.3) | 1 (1.9) | 4 (3.6) | 6 (3.4) |
| American Indian or Alaska Native | 0 | 1 (1.8) | 0 | 1 (0.9) | 1 (0.6) |
| Native Hawaiian or Pacific Islander | 0 | 1 (1.8) | 0 | 1 (0.9) | 1 (0.6) |
| Unknown | 2 (2.9) | 5 (8.8) | 2 (3.8) | 7 (6.4) | 9 (5.1) |
| Not reported | 6 (8.8) | 4 (7.0) | 3 (5.7) | 7 (6.4) | 13 (7.3) |
| Ethnicity, *n* (%) |  |  |  |  |  |
| Hispanic or Latino | 18 (26.5) | 18 (31.6) | 7 (13.2) | 25 (22.7) | 43 (24.2) |
| Not Hispanic or Latino | 47 (69.1) | 39 (68.4) | 43 (81.1) | 82 (74.5) | 129 (72.5) |
| Not reported | 3 (4.4) | 0 | 3 (5.7) | 3 (2.7) | 6 (3.4) |
| Weight, kg, mean (SD) | 89.40 (22.794) | 85.14 (22.412) | 91.71 (27.033) | 88.30 (24.852) | 88.72 (24.027) |
| Body-mass index, kg/m<sup>2</sup> |  |  |  |  |  |
| Mean (SD) | 30.94 (8.034) | 29.79 (6.684) | 31.20 (8.840) | 30.45 (7.772) | 30.64 (7.851) |
| ≥30, *n* (%) | 33 (48.5) | 22 (38.6) | 27 (50.9) | 49 (44.5) | 82 (46.1) |
| Days of COVID-19 illness prior to baseline, median (Q1:Q3) | 5.0 (4.0:8.0) | 7.0 (4.0:8.0) | 6.0 (4.0:7.0) | 6.0 (4.0:8.0) | 6.0 (4.0:8.0) |
| Baseline viral load |  |  |  |  |  |
| Median (Q1:Q3), log<sub>10</sub> copies/mL | 6.6 (5.1:7.7) | 6.5 (5.6:7.5) | 6.6 (5.6:7.7) | 6.5 (5.6:7.7) | 6.5 (5.6:7.7) |
| >10<sup>4</sup> copies/mL | 62 (91.2) | 53 (93.0) | 48 (90.6) | 101 (91.8) | 163 (91.6) |
| >10<sup>6</sup> copies/mL | 42 (61.8) | 38 (66.7) | 37 (69.8) | 75 (68.2) | 117 (65.7) |
| Concomitant medications, *n* (%) |  |  |  |  |  |
| Remdesivir | 40 (58.8) | 33 (57.9) | 39 (73.6) | 72 (65.5) | 112 (62.9) |
| Systemic corticosteroids | 50 (73.5) | 44 (77.2) | 47 (88.7) | 91 (82.7) | 141 (79.2) |
| Use of supplemental oxygen, *n* (%) | 53 (77.9) | 40 (70.2) | 34 (64.2) | 74 (67.3) | 127 (71.3) |
| Immunocompromised, *n* (%)<sup>b</sup> | 20 (29.4) | 11 (19.3) | 13 (24.5) | 24 (21.8) | 44 (24.7) |

| **Baseline neutralizing antibody status: Positive** | ***n* = 222** | ***n* = 213** | ***n* = 208** | ***n* = 421** | ***n* = 643** |
| --- | --- | --- | --- | --- | --- |
| Age, years |  |  |  |  |  |
| Median (range) | 60.0 (22:95) | 59.0 (20:90) | 60.5 (20:90) | 60.0 (20:90) | 60.0 (20:95) |
| ≥65, *n* (%) | 82 (36.9) | 70 (32.9) | 84 (40.4) | 154 (36.6) | 236 (36.7) |
| Male sex, *n* (%) | 123 (55.4) | 121 (56.8) | 119 (57.2) | 240 (57.0) | 363 (56.5) |
| Race, *n* (%) |  |  |  |  |  |
| White | 132 (59.5) | 136 (63.8) | 123 (59.1) | 259 (61.5) | 391 (60.8) |
| Black or African American | 30 (13.5) | 24 (11.3) | 29 (13.9) | 53 (12.6) | 83 (12.9) |
| Asian | 9 (4.1) | 12 (5.6) | 12 (5.8) | 24 (5.7) | 33 (5.1) |
| American Indian or Alaska Native | 8 (3.6) | 5 (2.3) | 12 (5.8) | 17 (4.0) | 25 (3.9) |
| Native Hawaiian or Pacific Islander | 2 (0.9) | 0 | 2 (1.0) | 2 (0.5) | 4 (0.6) |
| Unknown | 15 (6.8) | 13 (6.1) | 13 (6.3) | 26 (6.2) | 41 (6.4) |
| Not reported | 26 (11.7) | 23 (10.8) | 17 (8.2) | 40 (9.5) | 66 (10.3) |
| Ethnicity, *n* (%) |  |  |  |  |  |
| Hispanic or Latino | 90 (40.5) | 86 (40.4) | 80 (38.5) | 166 (39.4) | 256 (39.8) |
| Not Hispanic or Latino | 123 (55.4) | 117 (54.9) | 121 (58.2) | 238 (56.5) | 361 (56.1) |
| Not reported | 9 (4.1) | 10 (4.7) | 7 (3.4) | 17 (4.0) | 26 (4.0) |
| Weight, kg, mean (SD) | 89.13 (24.328) | 89.78 (24.988) | 90.33 (25.582) | 90.05 (25.256) | 89.73 (24.924) |
| Body-mass index, kg/m<sup>2</sup> |  |  |  |  |  |
| Mean (SD) | 31.62 (7.510) | 31.91 (8.112) | 31.72 (8.189) | 31.82 (8.141) | 31.75 (7.921) |
| ≥30, *n* (%) | 118 (53.2) | 106 (49.8) | 106 (51.0) | 212 (50.4) | 330 (51.3) |
| Days of COVID-19 illness prior to baseline, median (Q1:Q3) | 7.0 (5.0:8.0) | 7.0 (5.0:8.0) | 7.0 (5.0:8.0) | 7.0 (5.0:8.0) | 7.0 (5.0:8.0) |
| Baseline viral load |  |  |  |  |  |
| Median (Q1:Q3), log<sub>10</sub> copies/mL | 5.1 (4.3:6.1) | 5.1 (4.2:6.2) | 5.4 (4.2:6.4) | 5.2 (4.2:6.3) | 5.2 (4.2:6.2) |
| >10<sup>4</sup> copies/mL | 184 (82.9) | 171 (80.3) | 164 (78.8) | 335 (79.6) | 519 (80.7) |
| >10<sup>6</sup> copies/mL | 64 (28.8) | 62 (29.1) | 67 (32.2) | 129 (30.6) | 193 (30.0) |
| Concomitant medications, *n* (%) |  |  |  |  |  |
| Remdesivir | 139 (62.6) | 120 (56.3) | 125 (60.1) | 245 (58.2) | 384 (59.7) |
| Systemic corticosteroids | 177 (79.7) | 166 (77.9) | 181 (87.0) | 347 (82.4) | 524 (81.5) |
| Use of supplemental oxygen, *n* (%) | 177 (79.7) | 165 (77.5) | 175 (84.1) | 340 (80.8) | 517 (80.4) |
| Immunocompromised, *n* (%)<sup>b</sup> | 30 (13.5) | 32 (15.0) | 21 (10.1) | 53 (12.6) | 83 (12.9) |

<sup>a</sup>Seropositive mFAS presented.
<sup>b</sup>Immunocompromised patients include those with immunological diseases, are immunosuppressed, or have immunodeficiencies.
CAS+IMD, casirivimab and imdevimab; COVID-19, coronavirus disease 2019; IV, intravenous; mFAS, modified full analysis set; Q, quartile.

### Virologic efficacy

In seropositive patients on low-flow or no supplemental oxygen, treatment with CAS+IMD reduced viral load, relative to placebo, at all time points evaluated in patients who were negative or borderline for neutralizing antibodies (**Fig. 2A**) but not in patients with measurable neutralizing activity (**Fig. 2B**). In patients who were negative or borderline for neutralizing antibodies, a significant reduction in viral load with CAS+IMD versus placebo was observed as early as the first follow-up time point on day 3 and continued through day 11 (**Table S3**). Least-squares (LS) mean time-weighted average (TWA) daily change in viral load from baseline (day 1) through day 3 was –0.27 log_10_ copies/mL (95% confidence interval [CI]: –0.48 to –0.05) in the placebo group compared with –0.66 log_10_ copies/mL (95% CI: –0.83 to –0.48) in the CAS+IMD combined dose group, with an LS mean difference versus placebo of – 0.39 log_10_ copies/mL (95% CI: –0.66 to –0.11; nominal *P* = 0.0061). LS mean TWA daily change in viral load from baseline through day 11 was –1.33 log_10_ copies/mL (95% CI: –1.64 to –1.03) in the placebo group compared with –1.87 log_10_ copies/mL (95% CI: –2.11 to –1.63) in the CAS+IMD combined dose group, with an LS mean difference versus placebo of –0.54 log_10_ copies/mL (95% CI: –0.93 to –0.15; nominal *P* = 0.0067).

**FIG 2.**
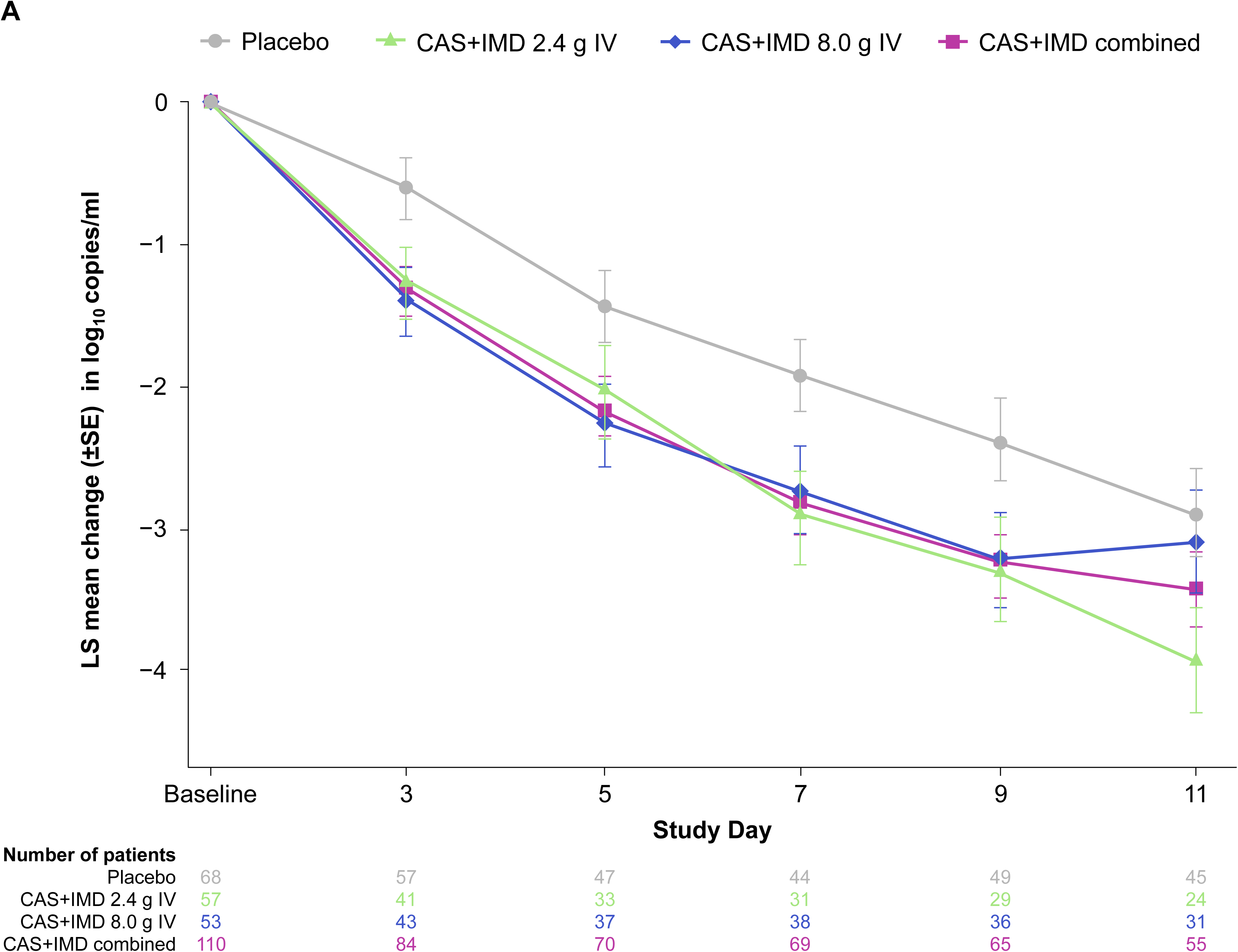
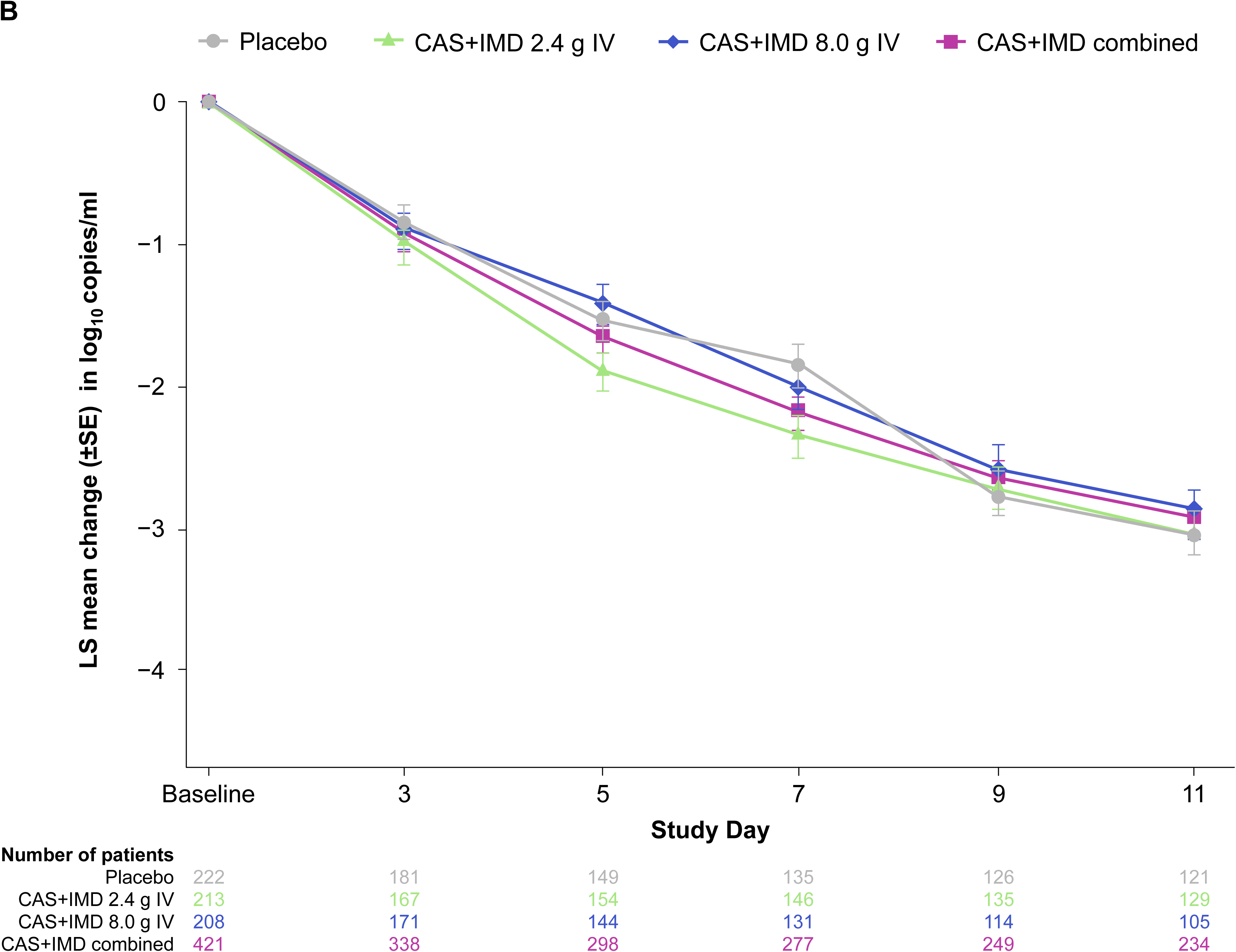
Change from baseline in viral load in seropositive patients by baseline neutralizing antibody status. (A) LS mean viral load following administration of CAS+IMD (2.4 g, 8.0 g, or combined analysis of 2.4 g and 8.0 g) or placebo for seropositive patients who were negative or borderline for neutralizing antibodies. (B) The same but for seropositive patients who were positive for neutralizing antibodies. Seropositive mFAS presented. CAS+IMD, casirivimab and imdevimab; IV, intravenous; mFAS, modified full analysis set; LS, least-squares.

### Clinical efficacy: death or mechanical ventilation

Though it was limited by small numbers, in seropositive patients on low-flow or no supplemental oxygen, a trend toward benefit in the proportion of patients who died or required mechanical ventilation was observed with CAS+IMD treatment versus placebo in patients who were negative or borderline for neutralizing antibodies (**Fig. 3A**). In this subset of patients, the proportion who died or required mechanical ventilation from days 1 to 29 was 19.1% (13/68) in the placebo group compared with 10.9% (12/110) in the CAS+IMD combined dose group (relative risk reduction, 49.2%; nominal *P* = 0.1125; **Table S4**).

**FIG 3.**
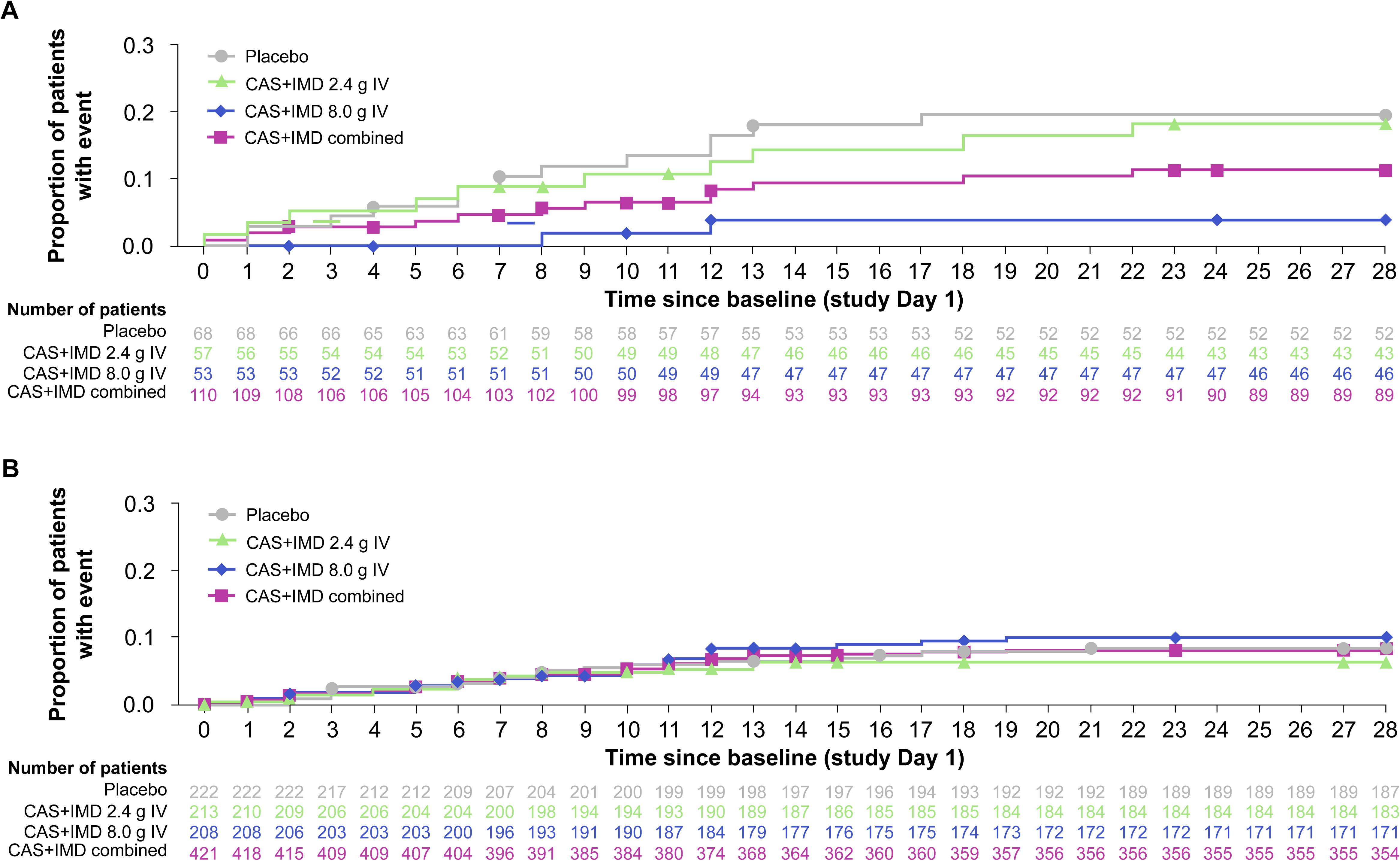
Cumulative incidence of death or mechanical ventilation in seropositive patients by baseline neutralizing antibody status. Kaplan–Meier curves for the proportion of patients who died or required mechanical ventilation through study day 29, after administration of CAS+IMD (2.4 g, 8.0 g, or combined analysis of 2.4 g and 8.0 g) or placebo in (A) patients who were negative or borderline for neutralizing antibodies or (B) patients who were positive for neutralizing antibodies. Symbols indicate censoring. Seropositive mFAS presented. CAS+IMD, casirivimab and imdevimab; IV, intravenous; mFAS, modified full analysis set.

In seropositive patients who were positive for neutralizing antibodies, no measurable benefit or harm in the proportion of patients who died or required mechanical ventilation was observed (**Fig. 3B**).

### Clinical efficacy: all-cause mortality

Though it was also limited by small numbers, in seropositive patients on low-flow or no supplemental oxygen, a trend toward benefit in all-cause mortality was observed with CAS+IMD treatment versus placebo in patients who were negative or borderline for neutralizing antibodies (**Fig. 4A**). In this subset of patients, the proportion of patients who died from days 1 to 29 was 16.2% (11/68) in the placebo group compared with 9.1% (10/110) in the CAS+IMD combined dose group (relative risk reduction, 43.8%; nominal *P* = 0.1190; **Table S4**).

**FIG 4.**
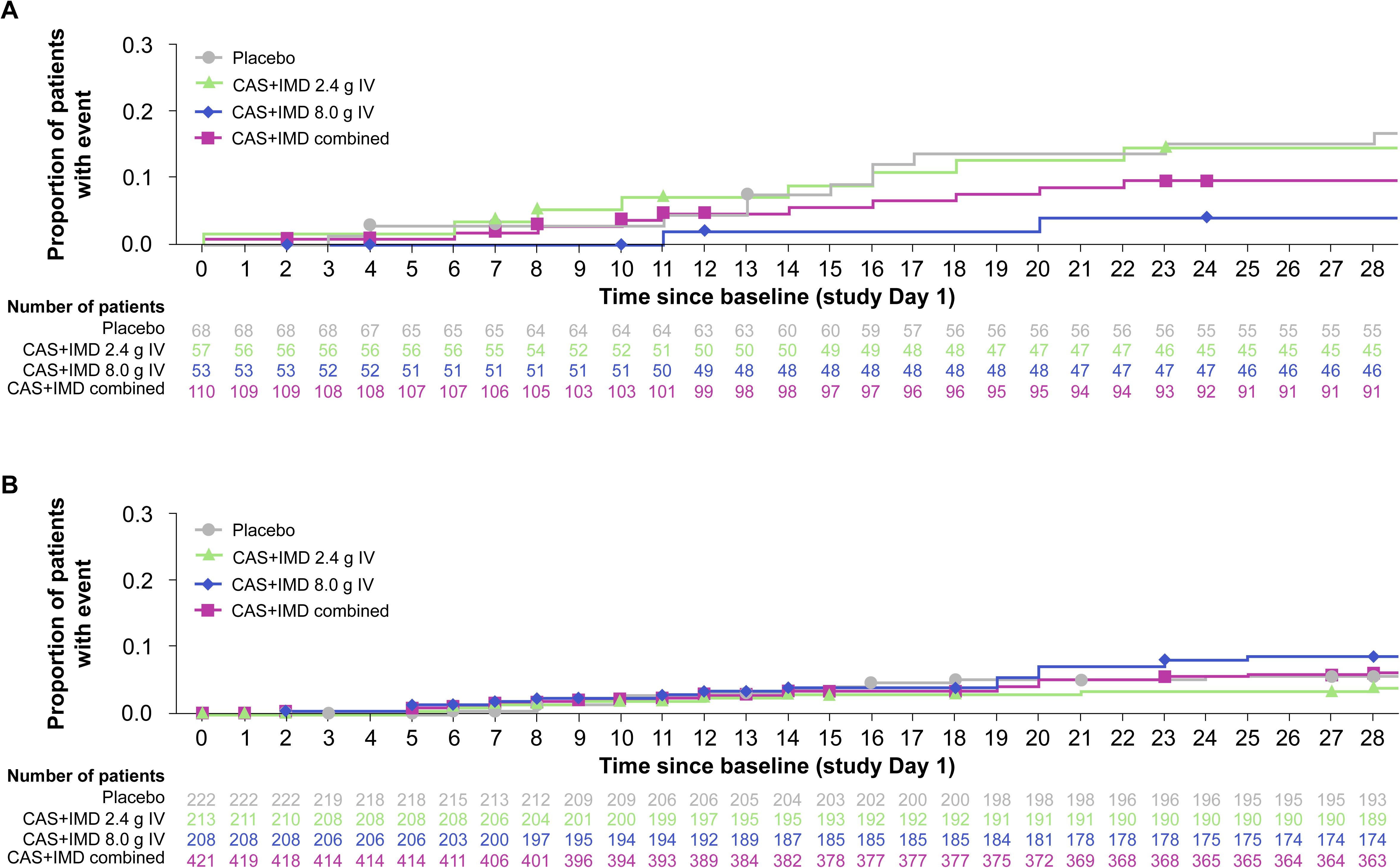
Cumulative incidence of death in seropositive patients by baseline neutralizing antibody status. Kaplan–Meier curves for the proportion of patients who died through study day 29, after administration of CAS+IMD (2.4 g, 8.0 g, or combined analysis of 2.4 g and 8.0 g) or placebo in (A) patients who were negative or borderline for neutralizing antibodies or (B) patients who were positive for neutralizing antibodies. Symbols indicate censoring. Seropositive mFAS presented. CAS+IMD, casirivimab and imdevimab; IV, intravenous; mFAS, modified full analysis set.

In seropositive patients who were positive for neutralizing antibodies, no measurable benefit or harm in all-cause mortality was observed (**Fig. 4B**).

### Safety

While event rates were small, in the subset of patients who were negative or borderline for neutralizing antibodies, serious adverse events (SAEs) were reported by 30.9% (21/68) and 29.1% (32/110) in the placebo and CAS+IMD combined dose groups, respectively (**Table 2**). In the subset of patients who were positive for neutralizing antibodies, SAEs were reported by 16.2% (36/222) and 14.7% (62/421) in the placebo and CAS+IMD combined dose groups, respectively.

**TABLE 2.**
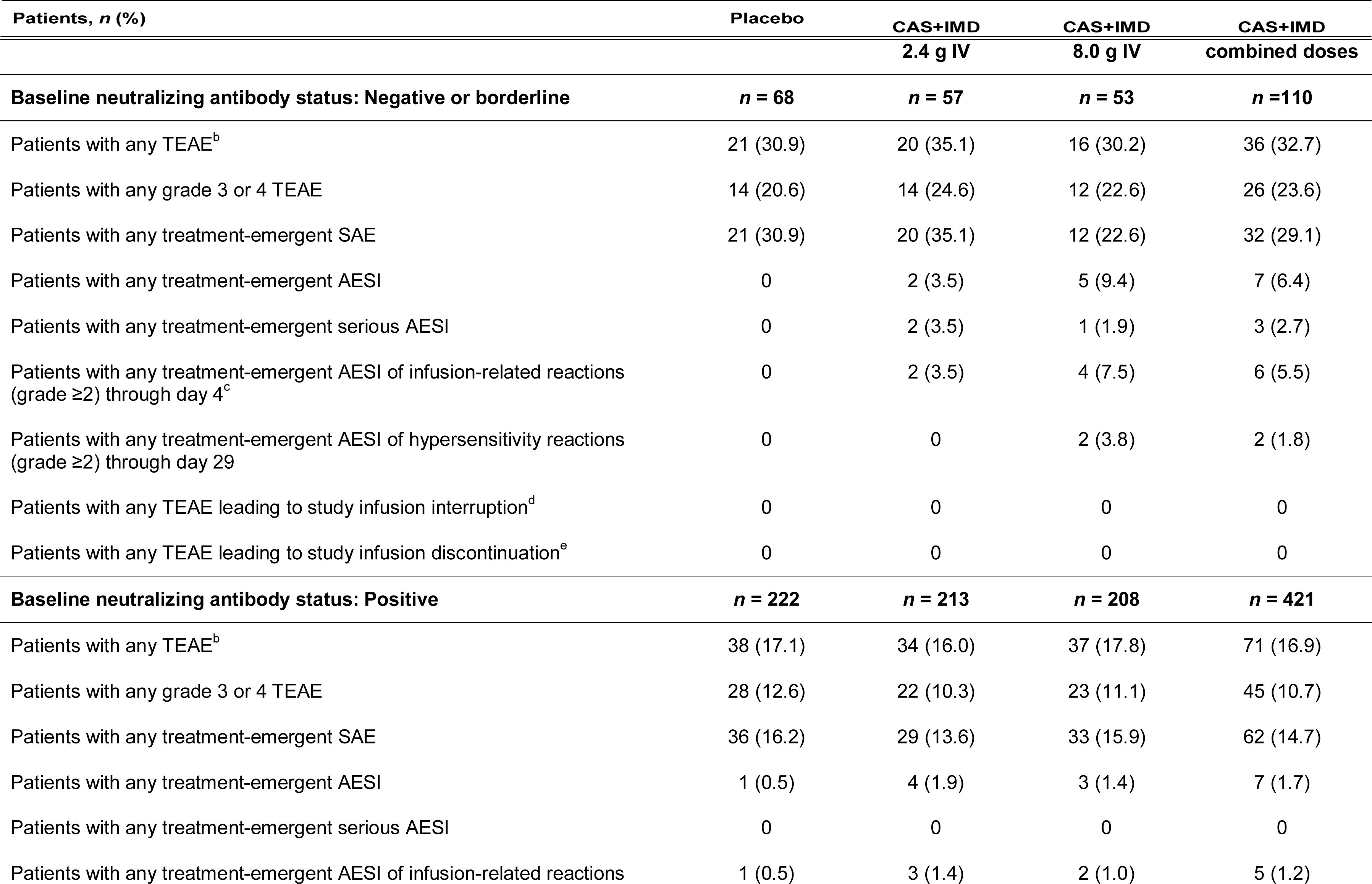
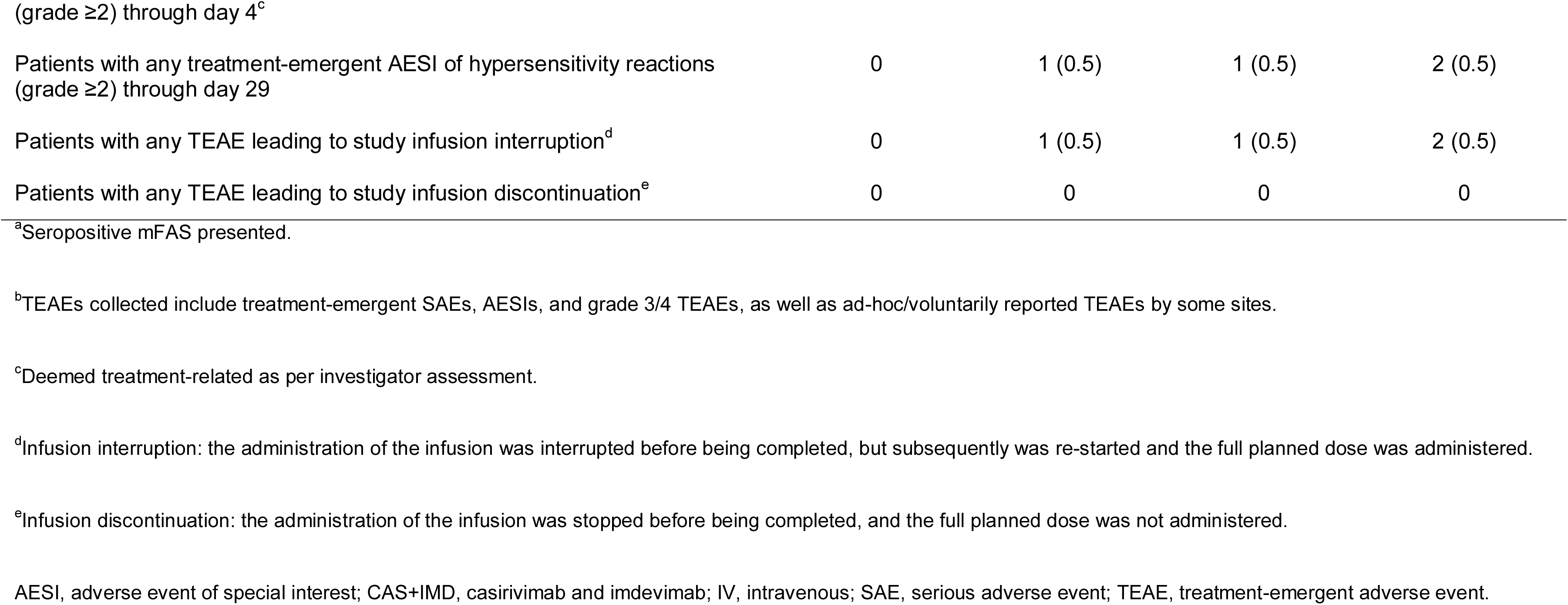
Overview of TEAEs in seropositive patients by baseline neutralizing antibody status^a^

| Patients, *n* (%) | Placebo | CAS+IMD | CAS+IMD | CAS+IMD |
| --- | --- | --- | --- | --- |

|  |  | 2.4 g IV | 8.0 g IV | combined doses |
| --- | --- | --- | --- | --- |
| **Baseline neutralizing antibody status: Negative or borderline** | **n = 68** | **n = 57** | **n = 53** | **n = 110** |
| Patients with any TEAE<sup>b</sup> | 21 (30.9) | 20 (35.1) | 16 (30.2) | 36 (32.7) |
| Patients with any grade 3 or 4 TEAE | 14 (20.6) | 14 (24.6) | 12 (22.6) | 26 (23.6) |
| Patients with any treatment-emergent SAE | 21 (30.9) | 20 (35.1) | 12 (22.6) | 32 (29.1) |
| Patients with any treatment-emergent AESI | 0 | 2 (3.5) | 5 (9.4) | 7 (6.4) |
| Patients with any treatment-emergent serious AESI | 0 | 2 (3.5) | 1 (1.9) | 3 (2.7) |
| Patients with any treatment-emergent AESI of infusion-related reactions (grade ≥2) through day 4<sup>c</sup> | 0 | 2 (3.5) | 4 (7.5) | 6 (5.5) |
| Patients with any treatment-emergent AESI of hypersensitivity reactions (grade ≥2) through day 29 | 0 | 0 | 2 (3.8) | 2 (1.8) |
| Patients with any TEAE leading to study infusion interruption<sup>d</sup> | 0 | 0 | 0 | 0 |
| Patients with any TEAE leading to study infusion discontinuation<sup>e</sup> | 0 | 0 | 0 | 0 |
| **Baseline neutralizing antibody status: Positive** | **n = 222** | **n = 213** | **n = 208** | **n = 421** |
| Patients with any TEAE<sup>b</sup> | 38 (17.1) | 34 (16.0) | 37 (17.8) | 71 (16.9) |
| Patients with any grade 3 or 4 TEAE | 28 (12.6) | 22 (10.3) | 23 (11.1) | 45 (10.7) |
| Patients with any treatment-emergent SAE | 36 (16.2) | 29 (13.6) | 33 (15.9) | 62 (14.7) |
| Patients with any treatment-emergent AESI | 1 (0.5) | 4 (1.9) | 3 (1.4) | 7 (1.7) |
| Patients with any treatment-emergent serious AESI | 0 | 0 | 0 | 0 |
| Patients with any treatment-emergent AESI of infusion-related reactions | 1 (0.5) | 3 (1.4) | 2 (1.0) | 5 (1.2) |
| Patients with any treatment-emergent AESI of hypersensitivity reactions (grade $\geq 2$) through day 29 | 0 | 1 (0.5) | 1 (0.5) | 2 (0.5) |
| Patients with any TEAE leading to study infusion interruption<sup>d</sup> | 0 | 1 (0.5) | 1 (0.5) | 2 (0.5) |
| Patients with any TEAE leading to study infusion discontinuation<sup>e</sup> | 0 | 0 | 0 | 0 |

In the subset of patients who were negative or borderline for neutralizing antibodies, more experienced adverse events that resulted in death in the placebo group compared with those in the CAS+IMD combined dose group (17.6% [12/68] placebo versus 10.0% [11/110] CAS+IMD; **Table S5**), consistent with the treatment benefit highlighted in the efficacy section. Interestingly, this trend was not observed in the subset of patients who were positive for neutralizing antibodies (6.3% [14/222] placebo versus 7.1% [30/421] CAS+IMD).

In the subset of patients who were negative or borderline for neutralizing antibodies, no adverse events of special interests (AESIs; grade ≥2 infusion-related reactions and grade ≥2 hypersensitivity reactions) were reported in the placebo group while AESIs were reported by 6.4% (7/110) of patients in the CAS+IMD combined dose group (**Table S6**). In the subset of patients who were positive for neutralizing antibodies, AESIs were reported by 0.5% (1/222) and 1.7% (7/421) in the placebo and CAS+IMD combined dose groups, respectively.

## DISCUSSION

The clinical benefit of CAS+IMD in hospitalized seronegative patients with COVID-19 was previously demonstrated in the RECOVERY study and the primary analysis of Study 2066 (11, 12). In these studies, no clinical benefit was observed in patients with baseline seropositivity to SARS-CoV-2 with active treatment relative to placebo. Presumably, the endogenous neutralizing activity in patients mimics the activity of CAS+IMD; thus, viral measures and clinical outcomes in seropositive patients treated with placebo is similar to those treated with CAS+IMD. We now extend the findings from Study 2066 by demonstrating that there is a subset of hospitalized patients who have detectable antibodies against SARS-CoV-2 (ie, seropositive), but have antibodies that are not functioning appropriately to neutralize SARS-CoV-2 (ie, those who were negative or borderline for neutralizing antibodies in assays that measure the capability of patient serum to neutralize recombinant VSV encoding the SARS-CoV-2 spike glycoprotein), who may benefit from treatment with anti-SARS-CoV-2 monoclonal antibody therapies.

In this post-hoc analysis of hospitalized patients with COVID-19 on low-flow or no supplemental oxygen, we found that approximately 20% of seropositive patients were negative or borderline for neutralizing antibodies to SARS-CoV-2. In this subset of patients, CAS+IMD significantly reduced viral load compared to placebo (TWA daily change from baseline to day 11: LS mean reduction, –0.54 log_10_ copies/mL; nominal *P* = 0.0067; **Table S3**). In contrast, a significant reduction in viral load was not observed relative to placebo in seropositive patients with measurable neutralizing activity. Furthermore, in the subset of seropositive patients who were negative or borderline for neutralizing antibodies, CAS+IMD treatment led to a trend towards benefit in death or mechanical ventilation (relative risk reduction, 42.9%; nominal *P* = 0.1125) as well mortality (relative risk reduction, 43.8%; nominal *P =* 0.1190) compared to placebo (**Table S4**).

In the subset of seropositive patients who were positive for neutralizing antibodies, CAS+IMD did not meaningfully impact death or mechanical ventilation (relative risk reduction, 0.4%; nominal *P* = 0.8689) or mortality (relative risk reduction, –9.9%; nominal *P* = 0.8893) compared to placebo, but no harm was observed. In contrast, a recent report of bamlanivimab treatment in hospitalized patients with COVID-19 from the ACTIV-3 study raised the question of whether that treatment might have caused harm in the patients who had already mounted an endogenous immune response to SARS-CoV-2 (17). In that analysis, when outcomes were analyzed by functional neutralizing antibody status, both death and the composite safety outcome (death, SAEs, organ failure, and serious co-infections) appeared to be worse in those who were positive for neutralizing antibodies.

While it is difficult to compare the findings from the ACTIV-3 study to the present analysis, given sample size limitations and differences in neutralization assays, both analyses indicate a clear trend toward benefit of monoclonal antibody therapy in patients lacking neutralizing antibody activity against SARS-CoV-2. The current study highlights that a subset of seropositive who lack neutralizing function against SARS-CoV-2 could still benefit from monoclonal antibody therapy.

Although the number of safety events was small, they were consistent with the primary analysis (12), and we did not observe any new or unknown safety signals or clustering in a particular treatment group and/or subset evaluated in this analysis. In the subset of seropositive patients who were negative or borderline for neutralizing antibodies, death rates were lower in the CAS+IMD group compared with the placebo group. This trend was not observed in the subset of seropositive patients who were positive for neutralizing antibodies at baseline. Finally, consistent with previous reports (12), we observed increased AESIs in the CAS+IMD group versus placebo in both subsets of seropositive patients.

As part of this analysis, we further characterized the serological status of seropositive patients, which was defined by a composite of three assays to detect antibodies against SARS-CoV-2, by looking at individual assay positivity as well as combinations of assay positivity (**Table S1 and Table S2**). While our data indicated that immunoglobulin (Ig) G assays may better differentiate neutralizing negative or borderline patients from neutralizing positive patients versus the composite of assays, there remain subsets of IgG seropositive patients lacking neutralizing activity who may benefit from treatment with CAS+IMD.

A key observation from this analysis is that utilizing serologic status (ie, seronegativity) to guide treatment decisions for patients with COVID-19 may fail to identify seropositive patients who may benefit from treatment with monoclonal antibodies, depending on the neutralizing potency of their endogenous antibodies. Moreover, functional antibodies generated from vaccination or natural infection with one variant of SARS-CoV-2 may not be effective against infection from a different variant. This clinical trial was conducted prior to the emergence of Omicron-lineage variants. While CAS+IMD is no longer in use in the US because of the predominance of Omicron (18), this analysis suggests that the next-generation of monoclonal antibodies for the treatment of COVID-19 may benefit patients whose endogenous antibodies lack anti-SARS-CoV-2 neutralizing activity, regardless of the overall baseline serostatus. Given that efficacy of COVID-19 vaccination wanes over time (1, 3, 19), the complexities of virus evolution (including regional differences), and the lack of available rapid point-of-care serology tests that reliably measure neutralizing function, identifying seropositive patients who would potentially benefit from treatment with monoclonal antibodies poses a challenge for patient care.

While we observed clear trends in improvement in both virologic and clinical endpoints in seropositive patients who were negative or borderline for neutralizing antibodies, this is a post-hoc analysis. Thus, all *P* values are considered nominal. Additionally, there was a relatively small number of patients in the seropositive negative or borderline subset (*N* = 178) relative to those with neutralizing activity, as well as compared to the number of seronegative patients, in this study.

Unlike the time of the pandemic during which Study 2066 was conducted, the majority of immunocompetent individuals are now expected to be seropositive for antibodies against SARS-CoV-2. The data presented here demonstrate that, in addition to the clinical benefit in seronegative hospitalized patients (11, 12), there may be a clinical benefit of anti-SARS-CoV-2 monoclonal antibody therapies in a subset of seropositive hospitalized patients who lack adequate neutralizing activity to the variant with which they are infected. Thus, further study of the potential of new monoclonal antibody therapies with activity to currently circulating SARS-CoV-2 variants in hospitalized patients, regardless of serostatus, is warranted.

## METHODS

### Trial design

The design of this adaptive, phase 1/2/3, double-blind, placebo-controlled trial to evaluate the efficacy, safety, and tolerability of CAS+IMD in hospitalized adult patients with COVID-19 (NCT04426695) has been previously described (12). Briefly, patients were enrolled in one of four cohorts based on disease severity: no supplemental oxygen (cohort 1A), low-flow oxygen (cohort 1), high-intensity oxygen (cohort 2), or mechanical ventilation (cohort 3). The trial proceeded through phase 2 for patients requiring no supplemental oxygen and phase 3 for patients requiring low-flow oxygen (O_2_ saturation >93% on low-flow oxygen via nasal cannula, simple face mask, or other similar device); together, these patients are the subject of this manuscript. For patients requiring high-intensity oxygen or mechanical ventilation, enrollment was paused early in the study per independent data monitoring committee (IDMC) recommendation as previously described (12), and these data are not included in this manuscript.

As previously described (12), patients were randomized 1:1:1 to a single intravenous dose of 2.4 g CAS+IMD, 8.0 g CAS+IMD, or placebo. The trial included a screening/baseline period, a hospitalization/post-discharge period (days 1 to 29), a monthly follow-up period, and an end-of-study visit (phase 1, day 169; phase 2/3, day 57).

### Patients

Patients were ≥18 years of age and hospitalized with confirmed SARS-CoV-2 within 72 hours of randomization and with symptom onset ≤10 days from randomization. Standard-of-care treatments for COVID-19 were permitted per the investigator. Full inclusion and exclusion criteria have been previously described (12). SARS-CoV-2 infection and baseline viral load were determined as previously described (Eurofins Viracor BioPharma Services, Inc., Lee’s Summit, MO, USA) (20).

### SARS-CoV-2 serostatus

All patients were assessed for the presence or absence of anti-SARS-CoV-2 antibodies at baseline by the following three assays comprising a composite serostatus: anti-spike [S1] IgA (EUROIMMUN), anti-spike [S1] IgG (EUROIMMUN), and anti-nucleocapsid IgG (Abbott). The composite of serology assays at baseline was run at a central laboratory (ICON Central Laboratories, Farmingdale, NY, USA). Patients underwent randomization regardless of their baseline serostatus, and were grouped for analyses as seropositive (if any baseline antibody test was positive) or seronegative (if all available baseline antibody tests were negative). Subjects who either had a borderline serostatus (if any test was borderline in the absence of any positive test result) or their test results were missing/not determined/pending were categorized as other.

### SARS-CoV-2 neutralization status

SARS-CoV-2 functional neutralizing titers were determined using a validated recombinant VSV neutralization assay where the VSV glycoprotein (G) was replaced by the SARS-CoV-2 spike (S) protein (utilizing the Wuhan sequence NC_045512.2) with a luciferase activity readout (Vyriad, Inc. and Imanis Life Sciences, LLC, Rochester, MN, USA) (16). Percent signals for each serum sample were determined from corrected raw light values. For analysis, patients were grouped as neutralizing positive (percent signal below the high positive control [HPC]), neutralizing negative (percent signal above low positive control [LPC]), neutralizing borderline (percent signal between HPC and LPC), or neutralizing indeterminant (percent signal for one replicate above LPC and one replicate below LPC).

### Outcome measures

In this post-hoc analysis, the following efficacy endpoints were evaluated in seropositive patients by neutralizing antibody status: 1) TWA daily change from baseline (day 1) in viral load in nasopharyngeal samples through day 11, 2) the proportion of patients who died or required mechanical ventilation from baseline (day 1) to day 29, and 3) the proportion of patients who died (all-cause mortality) from baseline (day 1) to day 29.

The following post-hoc safety endpoints were also evaluated in seropositive patients by neutralizing antibody status: the proportion of patients with 1) treatment-emergent SAEs through the end of the study, and 2) AESIs, specifically grade ≥2 infusion-related reactions through day 4 and grade ≥2 hypersensitivity reactions through day 29.

### Statistical analysis

As previously described (12), enrollment in this study was terminated on April 9, 2021 for strategic reasons, and not based on any safety concerns. Accordingly, enrollment of patients receiving low-flow (cohort 1) and no supplemental oxygen (cohort 1A) was prematurely terminated, but all ongoing patients were followed up through the end of the study. As such, phase 1, 2, and 3 patients on low-flow oxygen (cohort 1) were pooled with phase 2 patients with no supplemental oxygen (cohort 1A) for the current analysis. Additionally, it was elected to combine the CAS+IMD 2.4 g and 8.0 g dose groups for analysis.

The FAS includes all randomized patients who received any amount of study drug. The mFAS includes all FAS patients who had a positive central lab SARS-CoV-2 quantitative reverse transcriptase polymerase chain reaction result at baseline. The seropositive mFAS includes all patients in the mFAS who were grouped for analysis as seropositive, as described above; this population was used for all efficacy and safety analyses.

TWA daily change from baseline in viral load was analyzed using the analysis of covariance model, as previously described (12). The proportion of patients who died or required mechanical ventilation, as well as all-cause mortality, was analyzed using either the exact method for binomial distribution or asymptotic normal approximation method, as previously described (12). Safety endpoints were analyzed descriptively.

All reported *P* values for this post-hoc analysis are nominal. Missing data was handled as previously described (12).

### Trial oversight

Regeneron Pharmaceuticals, Inc. designed the trial and, with the trial investigators, gathered the data. Regeneron Pharmaceuticals, Inc. analyzed the data. The list of trial investigators has been previously described (12). The investigators, site personnel, and Regeneron Pharmaceuticals, Inc. were unaware of the treatment group assignments. An IDMC monitored unblinded data to make recommendations about trial modifications.

The trial was conducted in accordance with the principles of the Declaration of Helsinki, International Council for Harmonisation Good Clinical Practice guidelines, and applicable regulatory requirements. The local institutional review board or ethics committee at each study center oversaw trial conduct and documentation. Ethics approval for the COVID-19 Phase 2/3 Hospitalized Trial (COV-2066) was obtained from the following ethics review boards: Comissão de Ética para Análise de Projetos de Pesquisa do HCFMUSP, São Paulo, Brazil; Comitê De Ética Em Pesquisa Prof. Dr. Celso Figueirôa Hospital, Salvador, Bahia, Brazil; Comitê de Ética em Pesquisa da Faculdade de Medicina de Botucatu, São Paulo, Brazil; Comitê de Ética em Pesquisa da Universidade de Passo Fundo, Rio Grande do Sul, Brazil; Comitê de Ética em Pesquisa Envolvendo Seres Humanos da Universidade Comunitária da Servidão Anjo da Guarda, Santa Catarina, Brazil; Comitê de Ética em Pesquisa do Instituto Dante Pazzanese de Cardiologia, São Paulo, Brazil; Comitê de Ética em Pesquisa emSeres Humanos do Hospital das Clínicas da UFPR, Paraná, Brazil; Comitê de Ética em Pesquisa do Conjunto Hospitalar do Mandaqui, São Paulo, Brazil; Comité de ética de la Investigación de Clínica Las Condes, Santiago, Chile; Comité Ético Científico Facultad de Medicina Clínica Alemana Universidad del Desarrollo, Santiago, Chile; Comité de Ética en Investigación del Nuevo Hospital Civil de Guadalajara “Dr. Juan I Menchaca”, Guadalajara, Mexico; Comité de Ética en Investigación del Centro de Especialidades Médicas del Sureste, Merida, Yucatan, Mexico; Comité de Ética en Investigación del Hospital General de Culiacán, Culiacan, Mexico; Comité de ética en Investigación de Médica Sur, S.A.B de C.V., Ciudad de México, Mexico; Comité de Ética en Investigación del Hospital La Misión, Monterrey, Mexico; Hospital General de Occidente, Zapopan, Mexico; National Centre of Health Management, Chisinau, Republic of Moldova; Comisia Nationala de Bioetica a Medicamentului si Dispozitivelor Medicale, Bucuresti, Romania; Western Institutional Review Board, Puyallup, WA, USA; Providence St. Joseph Institutional Review Board, Renton, WA, USA; Tufts Health Sciences IRB, Boston, MA, USA; Providence Health and Services IRB, Portland, OR, USA; Lifespan IRB, Providence, RI, USA; St. Vincent Hospital and Health Care Center, Inc., Office of Clinical Trials, Indianapolis, IN, USA; BRANY, New Hyde Park, NY, USA; Institutional Review Board, University of Nebraska Medical Center, Omaha, NE, USA; Spectrum Health IRB, Grand Rapids, MI, USA; Research Compliance Office, Stanford University, Palo Alto, CA, USA; and Providence St. Joseph Health (PSJH) Institutional Review Board, Irvine, CA, USA. All patients provided written informed consent before participating in the trial.

## Supporting information

ICMJE form Yancopoulos

ICMJE form Geba

ICMJE form Hamilton

ICMJE form Mei

ICMJE form Dakin

ICMJE form Bhore

ICMJE form Ali

ICMJE form McCarthy

ICMJE form Somersan-Karakaya

ICMJE form Mahmood

ICMJE form Hooper

ICMJE form Weinreich

ICMJE form Mylonakis

ICMJE form Herman

## Data Availability

Qualified researchers may request access to study documents (including the clinical study report, study protocol with any amendments, blank case report form, and statistical analysis plan) that support the methods and findings reported in this manuscript. Individual anonymized participant data will be considered for sharing once the product and indication has been approved by major health authorities (eg, Food and Drug Administration, European Medicines Agency, Pharmaceuticals and Medical Devices Agency, etc.), if there is legal authority to share the data and there is not a reasonable likelihood of participant re-identification. Requests should be submitted to https://vivli.org/.

## ACKNOWLEDGEMENTS

We thank the patients who participated in this study, as well as their families; the study investigators; the members of the IDMC; Caryn Trbovic, PhD, from Regeneron Pharmaceuticals, Inc. for assistance with development of the manuscript; and Prime, Knutsford, UK, for formatting and copyediting suggestions.

## AUTHOR CONTRIBUTIONS

Conceptualisation: ATH, SS-K, SEM, SA, JM, RB, AM, GPG, PD, DMW, GDY, GAH, JDH

Data curation: ATH, SEM, JM, RB, JDH

Formal analysis: ATH, SS-K, SEM, SA, JM, RB, AM, GPG, PD, DMW, GDY, GAH, JDH

Investigation: EM

Methodology: ATH, SEM, JM, RB, GPG, JDH

Project administration: ATH, JDH

Resourcing: ATH, SS-K, SA, JDH

Software: JM, RB Visualisation: SEM, JM, RB

Validation: ATH, SEM, JM, RB, JDH

Supervision: JM, RB, AM, PD, DMW, GDY, GAH, JDH

Writing – original draft: ATH, SS-K, SEM, EM, JM, JDH

Writing – review and editing: ATH, SS-K, SEM, EM, SA, JM, RB, AM, GPG, PD, DMW, GDY, GAH, JDH

