## Supplementary material for "Casirivimab and Imdevimab Treatment in Seropositive, Hospitalized COVID-19 Patients With Non-neutralizing or Borderline Neutralizing Antibodies"

**SUPPLEMENTARY APPENDIX**

TABLE S1 Characterization of neutralization status in seropositive patients by individual serology assays^a^

| **Seroassay** | **patients, *n*** | **Seropositive patients positive for each assay, *n/N* (%)** | **Neutralization status, *n1/n* (%)** | | | | |
| --- | --- | --- | --- | --- | --- | --- | --- |
|  |  |  | **Positive** | **Negative** | **Borderline** | **Negative or borderline** | **U/M/I** |
| IgA spike | 764 | 764/1705^b^  (44.8) | 587/764 (76.8) | 107/764 (14.0) | 31/764 (4.1) | 138/764 (18.1) | 39/764 (5.1) |
| IgG spike | 372 | 372/1705^b^ (21.8) | 328/372 (88.2) | 23/372 (6.2) | 4/372 (1.1) | 27/372 (7.3) | 17/372 (4.6) |
| IgG NC | 534 | 534/1710^b^ (31.2) | 446/534 (83.5) | 44/534 (8.2) | 21/534 (3.9) | 65/534 (12.2) | 23/534 (4.3) |

^a^Seropositive mFAS presented.

^b^Denominators differ based on the number of patients with available data for each assay.

Ig, immunoglobulin; mFAS, modified full analysis set; NC, nucleocapsid; U/M/I, unknown/missing/indeterminate.

TABLE S2 Characterization of subgroup seropositivity by neutralization status^a^

| **Neutralizing status** | **Patients, *n*** | **Patients positive for each assay combination as presented, *n/N* (%)** | | | | | | | |
| --- | --- | --- | --- | --- | --- | --- | --- | --- | --- |
|  |  | **IgA spike^b^** | **IgG spike^b^** | **IgG NC^b^** | **IgA spike and  IgG spike^c^** | **IgA spike and  IgG NC^c^** | **IgG spike and  IgG NC^c^** | **IgG spike and/or IgG NC^d^** | **Triple positive^e^** |
| Negative or borderline | 178 | 91/178 (51.1) | 11/178 (6.2) | 27/178 (15.2) | 11/178 (6.2) | 33/178 (18.5) | 2/178 (1.1) | 40/178 (22.4) | 3/178 (1.7) |
| Negative | 138 | 75/138 (54.3) | 10/138 (7.2) | 19/138 (13.8) | 9/138 (6.5) | 21/138 (15.8) | 2/138 (1.4) | 31/138 (22.4) | 2/138 (1.4) |
| Borderline | 40 | 16/40 (40.0) | 1/40 (2.5) | 8/40 (20.0) | 2/40 (5.0) | 12/40 (30.0) | 0 (0) | 9/40 (22.5) | 1/40 (2.5) |
| Positive | 643 | 145/643 (22.6) | 8/643 (1.2) | 39/643 (6.1) | 44/643 (6.8) | 131/643 (20.4) | 9/643 (1.4) | 56/643 (8.6) | 267/643 (41.5) |

^a^Seropositive mFAS presented.

^b^Positive for only the individual assay specified.

^c^Positive for both of the assays specified but not positive for the remaining assay.

^d^Positive for either or both of the IgG assays (IgG spike, IgG NC) but not positive for IgA spike.

^e^Positive for all of the following three assays: IgA spike, IgG spike, and IgG NC.

Ig, immunoglobulin; mFAS, modified full analysis set; NC, nucleocapsid.

TABLE S3 Time-weighted average change in viral load from baseline in seropositive patients by baseline neutralizing antibody status^a,b^

|  | **Negative or borderline for neutralizing antibodies** | | | | **Positive for neutralizing antibodies** | | | |
| --- | --- | --- | --- | --- | --- | --- | --- | --- |
|  | **Placebo (*n* = 68)** | **CAS+IMD 2.4 g IV (*n* = 57)** | **CAS+IMD 8.0 g IV (*n* = 53)** | **CAS+IMD combined (*n* = 110)** | **Placebo (*n* = 222)** | **CAS+IMD 2.4 g IV (*n* = 213)** | **CAS+IMD 8.0 g IV (*n* = 208)** | **CAS+IMD combined (*n* = 421)** |
| Baseline (Day 1) to Day 3 |  |  |  |  |  |  |  |  |
| Patients, *n* | 57 | 41 | 43 | 84 | 181 | 167 | 171 | 338 |
| LS mean change (SE), log_10_ copies/mL | –0.27 (0.11) | –0.64 (0.13) | –0.67 (0.12) | –0.66 (0.09) | –0.43 (0.06) | –0.52 (0.06) | –0.46 (0.06) | –0.49 (0.04) |
| 95% CI | –0.48,  –0.05 | –0.89,  –0.39 | –0.92,  –0.42 | –0.83,  –0.48 | –0.55,  –0.31 | –0.65,  –0.40 | –0.58,  –0.33 | –0.58,  –0.40 |
| Difference versus placebo, log_10_ copies/mL |  |  |  |  |  |  |  |  |
| LS mean (SE) | – | –0.37 (0.17) | –0.40 (0.16) | –0.39 (0.14) | – | –0.09 (0.09) | –0.03 (0.09) | –0.06 (0.08) |
| 95% CI | – | –0.70,  –0.04 | –0.73,  –0.07 | –0.66,  –0.11 | – | –0.27, 0.08 | –0.20, 0.14 | –0.21, 0.09 |
| Nominal *P* value | – | **0.0270** | **0.0163** | **0.0061** | – | 0.2894 | 0.7425 | 0.4248 |
| Baseline (Day 1) to Day 5 |  |  |  |  |  |  |  |  |
| Patients, *n* | 61 | 44 | 47 | 91 | 195 | 186 | 182 | 368 |
| LS mean change (SE), log_10_ copies/mL | –0.52 (0.13) | –1.02 (0.15) | –1.12 (0.14) | –1.07 (0.10) | –0.72 (0.07) | –0.92 (0.07) | –0.73 (0.07) | –0.83 (0.05) |
| 95% CI | –0.77, –0.27 | –1.32, –0.73 | –1.40, –0.83 | –1.27, –0.87 | –0.86, –0.59 | –1.06, –0.78 | –0.87, –0.59 | –0.93, –0.73 |
| Difference versus placebo, log_10_ copies/mL | – |  |  |  |  |  |  |  |
| LS mean (SE) | – | –0.50 (0.20) | –0.60 (0.19) | –0.55 (0.16) | – | –0.20 (0.10) | –0.01 (0.10) | –0.10 (0.09) |
| 95% CI | – | –0.89, –0.11 | –0.97, –0.22 | –0.87, –0.23 | – | –0.39, 0.00 | –0.21, 0.19 | –0.27, 0.07 |
| Nominal *P* value | – | **0.0113** | **0.0021** | **0.0009** | – | 0.0530 | 0.9416 | 0.2412 |
| Baseline (Day 1) to Day 7 |  |  |  |  |  |  |  |  |
| Patients, *n* | 61 | 49 | 50 | 99 | 201 | 193 | 184 | 377 |
| LS mean change (SE), log_10_ copies/mL | –0.81 (0.13) | –1.32 (0.15) | –1.49 (0.15) | –1.40 (0.10) | –0.93 (0.07) | –1.23 (0.07) | –0.99 (0.07) | –1.11 (0.05) |
| 95% CI | –1.07, –0.55 | –1.61, –1.02 | –1.78, –1.20 | –1.61, –1.19 | –1.07, –0.79 | –1.37, –1.09 | –1.14, –0.85 | –1.22, –1.01 |
| Difference versus placebo, log_10_ copies/mL |  |  |  |  |  |  |  |  |
| LS mean (SE) | – | –0.51 (0.20) | –0.68 (0.20) | –0.59 (0.17) | – | –0.30 (0.10) | –0.06 (0.10) | –0.18 (0.09) |
| 95% CI | – | –0.90, –0.11 | –1.07, –0.29 | –0.92, –0.26 | – | –0.50, –0.09 | –0.26, 0.14 | –0.36, –0.01 |
| Nominal *P* value | – | **0.0123** | **0.0008** | **0.0006** | – | **0.0041** | **0.5592** | **0.0431** |
| Baseline (Day 1) to Day 9 |  |  |  |  |  |  |  |  |
| Patients, *n* | 64 | 51 | 50 | 101 | 202 | 193 | 187 | 380 |
| LS mean change (SE), log_10_ copies/mL | –1.11 (0.15) | –1.63 (0.16) | –1.76 (0.16) | –1.69 (0.12) | –1.18 (0.08) | –1.48 (0.08) | –1.23 (0.08) | –1.35 (0.06) |
| 95% CI | –1.40, –0.82 | –1.95, –1.31 | –2.09, –1.44 | –1.92, –1.47 | –1.33, –1.03 | –1.63, –1.32 | –1.39, –1.08 | –1.46, –1.25 |
| Difference versus placebo, log_10_ copies/mL |  |  |  |  |  |  |  |  |
| LS mean (SE) | – | –0.52 (0.22) | –0.65 (0.22) | –0.58 (0.19) | – | –0.30 (0.11) | –0.05 (0.11) | –0.18 (0.09) |
| 95% CI | – | –0.95, –0.09 | –1.08, –0.22 | –0.95, –0.22 | – | –0.51, –0.09 | –0.27, 0.16 | –0.36, 0.01 |
| Nominal *P* value | – | **0.0186** | **0.0035** | **0.0020** | – | 0.0059 | 0.6180 | 0.0580 |
| Baseline (Day 1) to Day 11 |  |  |  |  |  |  |  |  |
| Patients, *n* | 64 | 51 | 50 | 101 | 203 | 194 | 189 | 383 |
| LS mean change (SE), log_10_ copies/mL | –1.33 (0.15) | –1.83 (0.17) | –1.92 (0.17) | –1.87 (0.12) | –1.36 (0.08) | –1.66 (0.08) | –1.41 (0.08) | –1.54 (0.06) |
| 95% CI | –1.64, –1.03 | –2.17, –1.49 | –2.27, –1.58 | –2.11, –1.63 | –1.52, –1.20 | –1.82, –1.50 | –1.57, –1.25 | –1.65, –1.42 |
| Difference versus placebo, log_10_ copies/mL |  |  |  |  |  |  |  |  |
| LS mean (SE) | – | –0.49 (0.23) | –0.59 (0.23) | –0.54 (0.20) | – | –0.30 (0.11) | –0.05 (0.12) | –0.18 (0.10) |
| 95% CI | – | –0.95, –0.04 | –1.05, –0.13 | –0.93, –0.15 | – | –0.52, –0.07 | –0.28, 0.18 | –0.37, 0.02 |
| Nominal *P* value | – | **0.0346** | **0.0124** | **0.0067** | – | **0.0092** | 0.6605 | 0.0765 |

^a^Seropositive mFAS presented.

^b^Nominal *P* values >0.05 are in bold.

CAS+IMD, casirivimab and imdevimab; CI, confidence interval; IV, intravenous; LS, least squares; mFAS, modified full analysis set.

TABLE S4 Clinical outcomes in seropositive patients by baseline neutralizing antibody status^a^

|  | **Negative or borderline for neutralizing antibodies** | | | | **Positive for neutralizing antibodies** | | | |
| --- | --- | --- | --- | --- | --- | --- | --- | --- |
|  | **Placebo (*n* = 68)** | **CAS+IMD 2.4 g IV (*n* = 57)** | **CAS+IMD 8.0 g IV (*n* = 53)** | **CAS+IMD combined (*n* = 110)** | **Placebo (*n* = 222)** | **CAS+IMD 2.4 g IV (*n*= 213)** | **CAS+IMD 8.0 g IV (*n* = 208)** | **CAS+IMD combined (*n*= 421)** |
| Proportion of patients who died or went on mechanical ventilation from baseline (Day 1) to Day 29 | | | | | | | | |
| *n*/total *N* (%) | 13/68 (19.1) | 10/57 (17.5) | 2/53 (3.8) | 12/110 (10.9) | 18/222 (8.1) | 13/213 (6.1) | 21/208 (10.1) | 34/421 (8.1) |
| Relative risk reduction vs placebo, % | – | 8.2 | 80.3 | 42.9 | – | 24.7 | –24.5 | 0.4 |
| 95% CI, % | – | –93.4, 56.5 | 16.3, 95.3 | –17.7, 72.3 | – | –49.8, 62.2 | –127.0, 31.7 | –72.2, 42.4 |
| Nominal *P* value | – | 0.7568 | 0.0122 | 0.1125 | – | 0.3537 | 0.5455 | 0.8689 |
| Proportion of patients who died from baseline (Day 1) to Day 29 | | | | | | | | |
| *n*/total *N* (%) | 11/68 (16.2) | 8/57 (14.0) | 2/53 (3.8) | 10/110 (9.1) | 12/222 (5.4) | 8/213 (3.8) | 17/208 (8.2) | 25/421 (5.9) |
| Relative risk reduction vs placebo, % | – | 13.2 | 76.7 | 43.8 | – | 30.5 | –51.2 | –9.9 |
| 95% CI, % | – | –101.0, 62.5 | –0.8, 94.6 | –25.2, 74.8 | – | –66.6, 71.0 | –208.9, 26.0 | –114.4, 43.7 |
| Nominal *P* value | – | 0.5985 | 0.0377 | 0.1190 | – | 0.3409 | 0.2964 | 0.8893 |

^a^Seropositive mFAS presented.

CAS+IMD, casirivimab and imdevimab; CI, confidence interval; IV, intravenous; mFAS, modified full analysis set.

TABLE S5 Adverse events leading to death in seropositive patients by baseline neutralizing antibody status^a^

| **Primary system organ class**  **Preferred term** | **Placebo** | **CAS+IMD  2.4 g IV** | **CAS+IMD  8.0 g IV** | **CAS+IMD  combined doses** |
| --- | --- | --- | --- | --- |
| **Baseline neutralizing antibody status: Negative or borderline** | ***n* = 68** | ***n* = 57** | ***n* = 53** | ***n* = 110** |
| TEAEs leading to death, n | 12 | 9 | 2 | 11 |
| Patients with at least one TEAE leading to death, n (%) | 12 (17.6) | 9 (15.8) | 2 (3.8) | 11 (10.0) |
| Cardiac disorders, *n* (%) | 1 (1.5) | 3 (5.3) | 0 | 3 (2.7) |
| Cardiac arrest | 1 (1.5) | 1 (1.8) | 0 | 1 (0.9) |
| Cardiogenic shock | 0 | 1 (1.8) | 0 | 1 (0.9) |
| Ventricular tachycardia | 0 | 1 (1.8) | 0 | 1 (0.9) |
| Infections and infestations, *n* (%) | 6 (8.8) | 1 (1.8) | 2 (3.8) | 3 (2.7) |
| COVID-19 | 1 (1.5) | 1 (1.8) | 1 (1.9) | 2 (1.8) |
| COVID-19 pneumonia | 3 (4.4) | 0 | 1 (1.9) | 1 (0.9) |
| Pulmonary sepsis | 2 (2.9) | 0 | 0 | 0 |
| Respiratory,­ thoracic, and mediastinal disorders, n (%) | 3 (4.4) | 3 (5.3) | 0 | 3 (2.7) |
| Acute respiratory failure | 1 (1.5) | 3 (5.3) | 0 | 3 (2.7) |
| Idiopathic pulmonary fibrosis | 1 (1.5) | 0 | 0 | 0 |
| Respiratory failure | 1 (1.5) | 0 | 0 | 0 |
| General disorders and administration site conditions, *n* (%) | 2 (2.9) | 2 (3.5) | 0 | 2 (1.8) |
| Death | 1 (1.5) | 2 (3.5) | 0 | 2 (1.8) |
| Multiple organ dysfunction syndrome | 1 (1.5) | 0 | 0 | 0 |
| **Baseline neutralizing antibody status: Positive** | ***n* = 222** | ***n* = 213** | ***n* =*n*208** | ***n* = 421** |
| TEAEs leading to death, n | 14 | 11 | 20 | 31 |
| Patients with at least one TEAE leading to death, n (%) | 14 (6.3) | 11 (5.2) | 19 (9.1) | 30 (7.1) |
| Respiratory,­ thoracic, and mediastinal disorders, *n* (%) | 4 (1.8) | 4 (1.9) | 6 (2.9) | 10 (2.4) |
| Acute respiratory failure | 0 | 1 (0.5) | 3 (1.4) | 4 (1.0) |
| Respiratory failure | 2 (0.9) | 1 (0.5) | 3 (1.4) | 4 (1.0) |
| Pulmonary artery thrombosis | 0 | 1 (0.5) | 0 | 1 (0.2) |
| Pulmonary embolism | 0 | 1 (0.5) | 0 | 1 (0.2) |
| Hypoxia | 1 (0.5) | 0 | 0 | 0 |
| Pneumonitis | 1 (0.5) | 0 | 0 | 0 |
| General disorders and administration site conditions, *n* (%) | 1 (0.5) | 2 (0.9) | 6 (2.9) | 8 (1.9) |
| Multiple organ dysfunction syndrome | 1 (0.5) | 2 (0.9) | 6 (2.9) | 8 (1.9) |
| Infections and infestations, *n* (%) | 5 (2.3) | 2 (0.9) | 5 (2.4) | 7 (1.7) |
| COVID-19 | 3 (1.4) | 0 | 3 (1.4) | 3 (0.7) |
| COVID-19 pneumonia | 0 | 0 | 1 (0.5) | 1 (0.2) |
| Pneumonia | 0 | 0 | 1 (0.5) | 1 (0.2) |
| Septic shock | 1 (0.5) | 1 (0.5) | 0 | 1 (0.2) |
| Urosepsis | 0 | 1 (0.5) | 0 | 1 (0.2) |
| Pulmonary sepsis | 1 (0.5) | 0 | 0 | 0 |
| Cardiac disorders, *n* (%) | 3 (1.4) | 2 (0.9) | 3 (1.4) | 5 (1.2) |
| Cardio-respiratory arrest | 1 (0.5) | 1 (0.5) | 2 (1.0) | 3 (0.7) |
| Acute myocardial infarction | 0 | 0 | 1 (0.5) | 1 (0.2) |
| Cardiac arrest | 0 | 1 (0.5) | 0 | 1 (0.2) |
| Acute left ventricular failure | 1 (0.5) | 0 | 0 | 0 |
| Atrial fibrillation | 1 (0.5) | 0 | 0 | 0 |
| Nervous system disorders, *n* (%) | 1 (0.5) | 1 (0.5) | 0 | 1 (0.2) |
| Metabolic encephalopathy | 0 | 1 (0.5) | 0 | 1 (0.2) |
| Dementia Alzheimer's type | 1 (0.5) | 0 | 0 | 0 |

^a^Seropositive mFAS presented.

A patient who reported two or more adverse events with different preferred terms within the same system organ class is counted only once in that system organ class. A patient who reported two or more adverse events with the same preferred term is counted only once for that term.

Primary system organ classes are sorted according to decreasing order of frequency of all treatment groups combined. Within each system organ class, preferred terms are sorted by decreasing frequency.

CAS+IMD, casirivimab and imdevimab; COVID-19, coronavirus disease 2019; IV, intravenous; mFAS, modified full analysis set.

**TABLE S6** Adverse events of special interest in seropositive patients by baseline neutralizing antibody status^a^

| **Primary system organ class**  **Preferred term** | **Placebo** | **CAS+IMD  2.4 g IV** | **CAS+IMD  8.0 g IV** | **CAS+IMD  combined doses** |
| --- | --- | --- | --- | --- |
| **Baseline neutralizing antibody status: Negative or borderline** | ***n* = 68** | ***n* = 57** | ***n* = 53** | ***n*= 110** |
| Number of AESIs | 0 | 2 | 9 | 11 |
| Number of patients with at least one AESI (%) | 0 | 2 (3.5) | 5 (9.4) | 7 (6.4) |
| General disorders and administration site conditions, *n* (%) | 0 | 1 (1.8) | 3 (5.7) | 4 (3.6) |
| Chills | 0 | 0 | 2 (3.8) | 2 (1.8) |
| Edema | 0 | 0 | 1 (1.9) | 1 (0.9) |
| Systemic inflammatory response syndrome | 0 | 1 (1.8) | 0 | 1 (0.9) |
| Respiratory,­ thoracic, and mediastinal disorders, *n* (%) | 0 | 1 (1.8) | 1 (1.9) | 2 (1.8) |
| Hypoxia | 0 | 1 (1.8) | 1 (1.9) | 2 (1.8) |
| Cardiac disorders, *n* (%) | 0 | 0 | 1 (1.9) | 1 (0.9) |
| Tachycardia | 0 | 0 | 1 (1.9) | 1 (0.9) |
| Gastrointestinal disorders, *n* (%) | 0 | 0 | 1 (1.9) | 1 (0.9) |
| Nausea | 0 | 0 | 1 (1.9) | 1 (0.9) |
| Vomiting | 0 | 0 | 1 (1.9) | 1 (0.9) |
| Immune system disorders, *n* (%) | 0 | 0 | 1 (1.9) | 1 (0.9) |
| Anaphylactic reaction | 0 | 0 | 1 (1.9) | 1 (0.9) |
| Nervous system disorders, *n* (%) | 0 | 0 | 1 (1.9) | 1 (0.9) |
| Headache | 0 | 0 | 1 (1.9) | 1 (0.9) |
| **Baseline neutralizing antibody status: Positive** | ***n* = 222** | ***n* = 213** | ***n* =208** | ***n* = 421** |
| Number of AESIs | 1 | 5 | 3 | 8 |
| Number of patients with at least one AESI (%) | 1 (0.5) | 4 (1.9) | 3 (1.4) | 7 (1.7) |
| Nervous system disorders, *n* (%) | 0 | 1 (0.5) | 1 (0.5) | 2 (0.5) |
| Dizziness | 0 | 0 | 1 (0.5) | 1 (0.2) |
| Hypoesthesia | 0 | 1 (0.5) | 0 | 1 (0.2) |
| Paresthesia | 0 | 1 (0.5) | 0 | 1 (0.2) |
| Respiratory,­ thoracic, and mediastinal disorders, *n* (%) | 0 | 1 (0.5) | 1 (0.5) | 2 (0.5) |
| Dyspnea | 0 | 0 | 1 (0.5) | 1 (0.2) |
| Hypoxia | 0 | 1 (0.5) | 0 | 1 (0.2) |
| General disorders and administration site conditions, *n* (%) | 0 | 0 | 1 (0.5) | 1 (0.2) |
| Infusion site pain | 0 | 0 | 1 (0.5) | 1 (0.2) |
| Investigations, *n* (%) | 0 | 1 (0.5) | 0 | 1 (0.2) |
| Aspartate aminotransferase increased | 0 | 1 (0.5) | 0 | 1 (0.2) |
| Skin and subcutaneous tissue disorders, *n* (%) | 0 | 1 (0.5) | 0 | 1 (0.2) |
| Pruritus | 0 | 1 (0.5) | 0 | 1 (0.2) |
| Injury,­ poisoning, and procedural complications, *n* (%) | 1 (0.5) | 0 | 0 | 0 |
| Infusion-related reaction | 1 (0.5) | 0 | 0 | 0 |

^a^Seropositive mFAS presented.

A patient who reported two or more adverse events with different preferred terms within the same system organ class is counted only once in that system organ class. A patient who reported two or more adverse events with the same preferred term is counted only once for that term.

Primary system organ classes are sorted according to decreasing order of frequency of all treatment groups combined. Within each system organ class, preferred terms are sorted by decreasing frequency.

AESI, adverse event of special interest; CAS+IMD, casirivimab and imdevimab; IV, intravenous; mFAS, modified full analysis set.

FIG S1 Flow diagram for the phase 1/2/3 population receiving low-flow or no supplemental oxygen

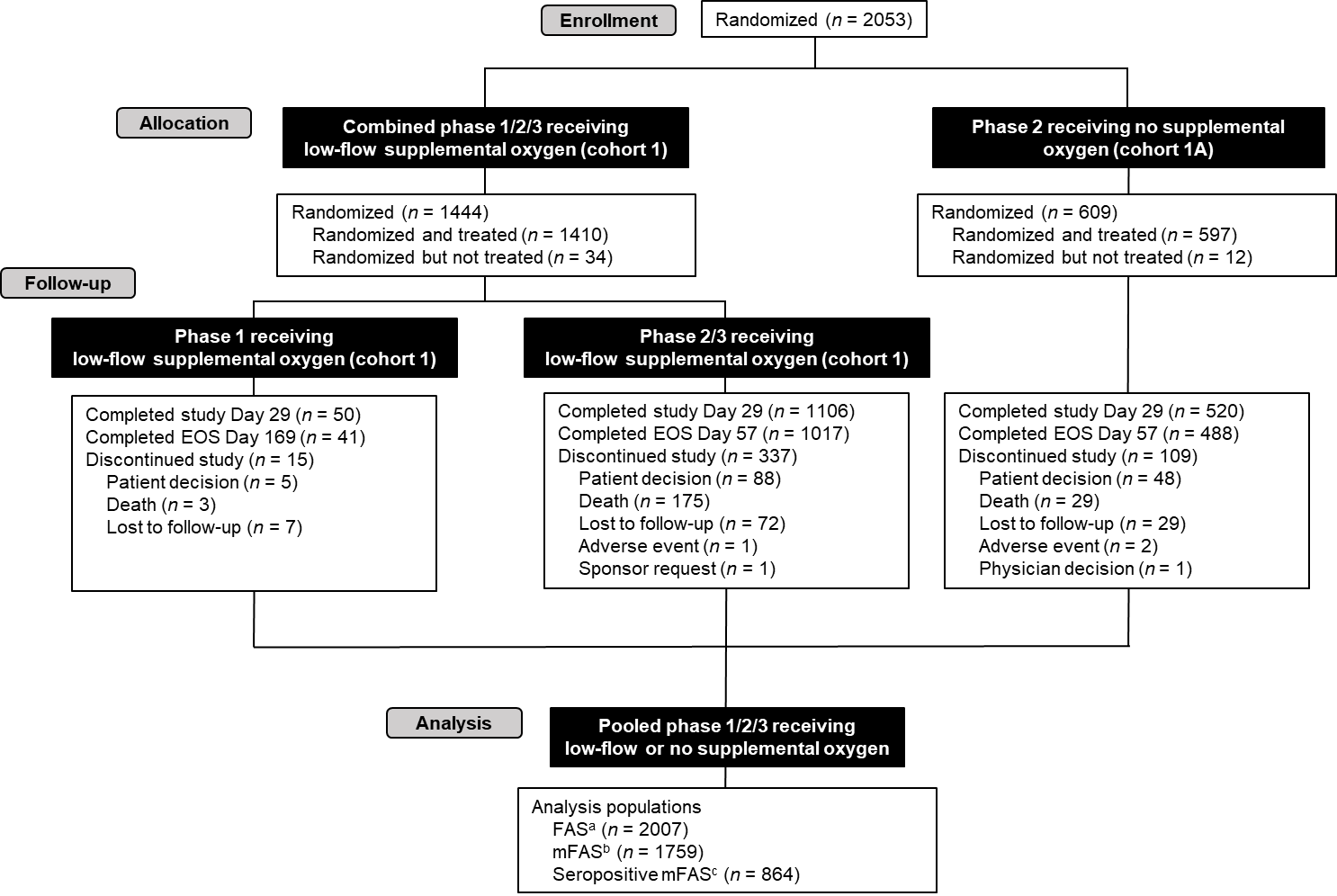

^a^The FAS includes all randomized patients who received at least one dose (full or partial) of the study drug. Analysis of the FAS population was according to the treatment allocated (as randomized).

^b^The mFAS includes all FAS patients with a positive SARS-CoV-2 RT-qPCR conducted in the central laboratory in nasopharyngeal swab samples at randomization, and analysis is based on the treatment allocated (as randomized).

^c^The seropositive mFAS is defined as all patients in mFAS in whom any baseline serology test (anti-spike [S1] IgA, anti-spike [S1] IgG, or anti-nucleocapsid IgG) was positive.

EOS, end of study; FAS, full analysis set; Ig, immunoglobulin; mFAS, modified full analysis set; RT-qPCR, quantitative reverse transcription polymerase chain reaction; SARS-CoV-2, severe acute respiratory syndrome coronavirus 2.
